# Large-scale social media language analysis reveals emotions and behaviours associated with nonmedical prescription drug use

**DOI:** 10.1101/2021.09.20.21263856

**Authors:** Mohammed Ali Al-Garadi, Yuan-Chi Yang, Yuting Guo, Sangmi Kim, Jennifer S. Love, Jeanmarie Perrone, Abeed Sarker

## Abstract

Nonmedical use of prescription drugs (NMPDU) is a global health concern. The extent of, behaviors and emotions associated with, and reasons for NMPDU are not well-captured through traditional instruments such as surveys, prescribing databases and insurance claims. Therefore, this study analyses ∼130 million public posts from 87,718 Twitter users in terms of expressed emotions, sentiments, concerns, and potential reasons for NMPDU via natural language processing. Our results show that users in the NMPDU group express more negative emotions and less positive emotions, more concerns about family, the past and body, and less concerns related to work, leisure, home, money, religion, health and achievement, compared to a control group (i.e., users who never reported NMPDU). NMPDU posts tend to be highly polarized, indicating potential emotional triggers. Gender-specific analysis shows that female users in the NMPDU group express more content related to positive emotions, anticipation, sadness, joy, concerns about family, friends, home, health and the past, and less about anger, compared to males. The findings of the study can enrich our understanding of NMPDU.

---

Nonmedical prescription drug use (NMPDU) involves the use of prescription drugs without a prescription or for reasons other than what the drug was intended for by the prescriber ^1^. NMPDU is an unremitting public health concern globally and in the United States (US) ^2^. Commonly misused prescription drugs include but are not limited to opioids, central nervous system stimulants, and benzodiazepines^3,4^. Increases in NMPDU over recent years have led to increased adverse health outcomes, including emergency department visits and overdose deaths^5^. In the US, more than 90,000 drug overdose deaths were recorded in 2020, many of which were caused by prescription drugs, often due to co-ingestion or polysubstance use^6,7^. While studies have attempted to characterize the reasons for NMPDU^8,9^, little is known about the emotional status of the consumers at the time of NMPDU. Studies investigating the influence of NMPDU on mental health have been primarily conducted through surveys. NMPDU involving opioids have been shown to be strongly associated with psychiatric disorders^10^ (data for the study was derived from the National Epidemiologic Survey on Alcohol and Related Conditions-III^10^). Analysis of data from the National Survey on Drug Use and Health (NSDUH) revealed associations between opioid misuse and suicide-related risk factors and that users involved in NMPDU of opioids being at higher risk of suicidality and suicidal ideation^11,12^ compared to those who never used these medications nonmedically. Past studies^3,13^ supported similar findings and showed associations between NMPDU of opioids and major depressive disorder or depressive symptoms.

Survey-based studies about NMPDU face several obstacles related to data collection, such as slow collection rates, high costs, and limited sample sizes. Importantly, studies using surveys are unable to capture naturally occurring emotions due to experimental or instrumental manipulations that could introduce measurement and observation biases^14^. Social media can address some of the shortcomings of such traditional survey-based studies. Social media presents a unique opportunity to collect information related to NMPDU for analysis at a large scale discreetly and unobtrusively so that the users’ expressions are not manipulated by experimental settings or processes. Also, the rising popularity of social media platforms has resulted in tremendous growth in the public sharing of information. Publicly available, user-generated social media data contain naturally-occurring communication phenomena describing users’ daily activities, issues, and concerns, which enable the execution of observational studies to understand social dynamics^15-17^ and human behaviours at the macro level, including behaviours related to NMPDU^18^. Indeed, past research has shown that social media users often share information about NMPDU publicly, which can be used for making macro-level assessments of drug abuse/misuse-related behaviors^19-21^. Recent studies^21-23^ validated the utility of social media as a platform for monitoring NMPDU. For instance, a qualitative assessment of the text content from Twitter on NMPDU (prescription opioid) delivered insights about the epidemic of use and misuse of PMs at specific times^22^. Multiple studies have suggested that although users engaging in NMPDU may not voluntarily report their nonmedical use to medical experts, their self-reports in social media are detectable^21,24,25^, and these can potentially be used for public health surveillance. A critical review^18^ concluded that social media big data could be an effective resource to comprehend, monitor, and intervene in drug misuses and addiction problems.

In addition to behaviours, emotion-related content in social media provides important information on the users’ psychological and physical health^26^. Negative emotion words of higher magnitudes are associated with greater psychological distress and worse physical health, while high-magnitude positive emotion words are associated with higher well-being and better physical health^26^. Demographic information about users, such as gender, may also be inferred from social media for differential behaviour analysis. For example, social media based research has shown that males and females have differing emotional tendencies under different circumstances, and certain online activities of female users are more susceptible to emotional orientations^27^. Recognizing the gender differences in user behaviours is a significant factor in user modelling, human-computer interaction, and the differences were investigated in previous studies through the analyses of lexical contents, including emoticons^28-30^. In the context of NMPDU, understanding gender differences between NMPDU users is particularly critical, as women specifically had often been underrepresented in past studies on the topic^31^.

In this study, we sought to employ natural language processing (NLP) and machine learning approaches to study a large dataset from Twitter about three common NMPDU categories and their combinations (opioids, benzodiazepines, stimulants, and polysubstance—misuse of two or more different NMPDU category at the same time, typically referred to as co-ingestion) to investigate and answer the following main research questions: (1) How do the emotional contents expressed in NMPDU groups’ Twitter profiles differ from those expressed in non-NMPDU (control group) groups’ Twitter profiles? (2) How do NMPDU tweets sentimentally differ from non-NMPDU tweets? And (3) how do personal, social, biological, and core drive concerns expressed in NMPDU groups’ Twitter profiles differ from those expressed in non-NMPDU groups’ Twitter profiles? In addition to attempting to answer these questions, we use topic modeling on NMPDU tweets to extract potential reasons for nonmedical uses of each category of drugs, and we compare the distributions (of all the variables mentioned above) across males and females.

## Results

### NMPDU (experimental group) and non-NMPDU (control group) users

We included a total of 87,718 Twitter users and their > 130 million posts in this study. To automatically characterize tweets (i.e., whether a tweet expresses self-reported NMPDU or not) mentioning specific medication keywords (see Supplementary S.2.1), we applied an automatic machine learning classifier, which was trained using a state-of-the-art NLP algorithm and a large manually-annotated dataset (see Methods). We then extracted all the publicly available posts (timelines) for all users who have posted at least one tweet classified as NMPDU. To build the non-NMPDU (control group) data, we randomly extracted publicly available timelines of users who may or may not have mentioned the medication keywords but whose posts were not classified as NMPDU. We excluded users with less than 500 publicly available tweets. Table 1 presents the distribution of users and tweets in the NMPDU and non-NMPDU groups.

**Table 1.** Number of unique users and tweets in both NMPDU and Non-NMPDU

|  | Unique users | Number of tweets |
| --- | --- | --- |
| **Opioids** | 8,290 | 8,084,203 |
| **Stimulants** | 18,540 | 39,917,674 |
| **Benzodiazepines** | 14,773 | 23,085,707 |
| **Polysubstance** | 8,230 | 11,164,212 |
| **Total number of unique users and number of tweets** |  |  |
| **Total NMPDU** | 49,833 | 82,251,796 |
| **Total non-NMPDU** | 37,885 | 55,352,960 |
| **Total** | 87,718 | 137,604,756 |

### Emotion analysis

We investigated the emotion content differences in users’ tweets from the NMPDU and non-NMPDU groups (Table 2). We performed linguistic emotion analysis of the complete profile contents for both groups using the lexicon curated by the National Research Council (NRC), Canada, which contains a comprehensive list of approximately 14,182 English words related to anger, fear, anticipation, trust, surprise, sadness, joy, sentiment (negative and positive), and disgust^32^. We then used the Anderson–Darling (AD) test^33^ and performed histogram analysis to check the normality distribution of the emotion-indicating variables in both groups. The histogram analysis and AD test results (see Supplementary S.3) confirmed the absence of normality distribution in all the emotion-indicating variables of both groups. Therefore, we used a nonparametric approach, the Mann–Whitney test^34,35^, to compare the distributions of emotion-indicating variables between users in the NMPDU and non-NMPDU groups. Table 2 presents the median Mann–Whitney U test results and the effect sizes of the comparisons between the NMPDU and control groups. It also presents comparisons between the NMPDU group and the control group for each medication category (i.e., opioids, benzodiazepines, stimulants, and polysubstance) in Supplementary S.4.

**Table 2.** Comparison of emotional differences between users from the NMPDU and control groups

| Emotions | Median: NMPDU group | Median: (Control group) | Different (NMPDU versus Control) Mann–Whitney U test results |  |
| --- | --- | --- | --- | --- |
|  |  |  | P-value (p) | Effect sizes (r) |
| **Positive** | 0.534 | 0.696 | <0.001 | 0.40** |
| **Trust** | 0.326 | 0.393 | <0.001 | 0.28* |
| **Anticipation** | 0.298 | 0.396 | <0.001 | 0.44** |
| **Fear** | 0.218 | 0.176 | <0.001 | 0.31** |
| **Anger** | 0.234 | 0.148 | <0.001 | 0.56*** |
| **Negative** | 0.458 | 0.337 | <0.001 | 0.45** |
| **Sadness** | 0.215 | 0.169 | <0.001 | 0.36** |
| **Joy** | 0.284 | 0.368 | <0.001 | 0.38** |
| **Surprise** | 0.145 | 0.190 | <0.001 | 0.39** |
| **Disgust** | 0.196 | 0.113 | <0.001 | 0.60*** |

Size effects(r): \*Small difference ( $0.1 \leq r < 0.3$ ). \*\*Moderate difference ( $0.3 \leq r < 0.5$ ). \*\*\*Large difference ( $0.5 \leq r < 0.7$ ).
\*\*\*\*Very large difference ( $r \geq 0.7$ )<sup>36</sup>

The users from the NMPDU group tend to share significantly more content related to fear (p < 0.001, r = 0.31), anger (p < 0.001, r = 0.56), negative emotion (p < 0.001, r = 0.45), sadness (p < 0.001, r = 0.36), and disgust (p < 0.001, r = 0.60) compared to the users from the control group. The NMPDU users share significantly less content related to positive emotions (p < 0.001, r = 0.40), joy (p < 0.001, r = 0.38), trust (p < 0.001, r = 0.28), anticipation (p < 0.001, r = 0.44), and surprise (p < 0.001, r = 0.39) than the users from the control group (Table 2). These findings are consistent across all four medication categories considered in this study (Supplementary S.4, Table 4).

### Gender differences in emotions within the NMPDU group

Within the NMPDU group, female users use more emotional content words/descriptors in the NMPDU-related social media posts as compared to male users (Table 3, Figure 2A). Specifically, female users express more content related to positive emotion (p < 0.001, r = 0.246), anticipation (p < 0.001, r = 0.247), sadness (p < 0.001, r = 0.21), joy (p < 0.001, r = 0.38) compared to male users. In contrast, male users express significantly more content related to anger (p < 0.001, r = 0.07) than female users. The results also show no significant difference between males and females in content related to trust, fear, surprise, disgust, and negative emotions.

**Table 3.** Comparison of emotions between male and female users from the NMPDU.

| Emotions | Median: Male NMPDU group | Median: Female NMPDU group | Different (Male NMPDU versus Female NMPDU )<br>Mann–Whitney U test results |  |
| --- | --- | --- | --- | --- |
|  |  |  | P-value (p) | Effect sizes (r) |
| **Positive** | 0.491 | 0.571 | <0.001 | 0.246* |
| **Trust** | 0.304 | 0.342 | >0.001 | - |
| **Anticipation** | 0.274 | 0.319 | <0.001 | 0.247* |
| **Fear** | 0.212 | 0.223 | >0.001 | - |
| **Anger** | 0.240 | 0.229 | <0.001 | 0.07* |
| **Negative** | 0.450 | 0.465 | >0.001 | - |
| **Sadness** | 0.202 | 0.229 | <0.001 | 0.21* |
| **Joy** | 0.249 | 0.317 | <0.001 | 0.38** |
| **Surprise** | 0.136 | 0.152 | >0.001 | - |
| **Disgust** | 0.195 | 0.198 | >0.001 | - |

Size effects(r): \*Small difference ( $0.1 \leq r < 0.3$ ). \*\*Medium difference ( $0.3 \leq r < 0.5$ ). \*\*\*Large difference ( $0.5 \leq r < 0.7$ ).
\*\*\*\*Very large difference ( $r \geq 0.7$ )<sup>36</sup>

### Sentiment strengths of NMPDU tweets

Sentiment analysis is a category of NLP methods that attempt to quantify positive or negative sentiments in text. We intended to measure and compare the sentiment polarities and strengths between the NMPDU and non-NMPDU tweets from the same users. As illustrated in Figure 1A, the NMPDU tweets contain larger magnitudes of extreme positive and negative sentiments (tweets with a positive score > 0.5 or a negative score < −0.5) compared to the non-NMPDU tweets. We empirically compared the sentiment strength means and confidence intervals for highly polarized tweets (sentiment < -0.5 or > 0.5) from the NMPDU and non-NMPDU categories Figure 1B. The sentiment strength means of the NMPDU tweets (both positive 95% CI [0.705, 0.711] and negative tweets 95% CI [-0.688, -0.680]) are higher in magnitude than the non-NMPDU tweets (positive 95% CI [0.652, 0.653] and negative tweets 95% CI [-0.637, -0.636]). The highly polarized nature of the NMPDU tweets indicate potential emotional triggers associated with NMPDU behaviour.

**Figure 1.**
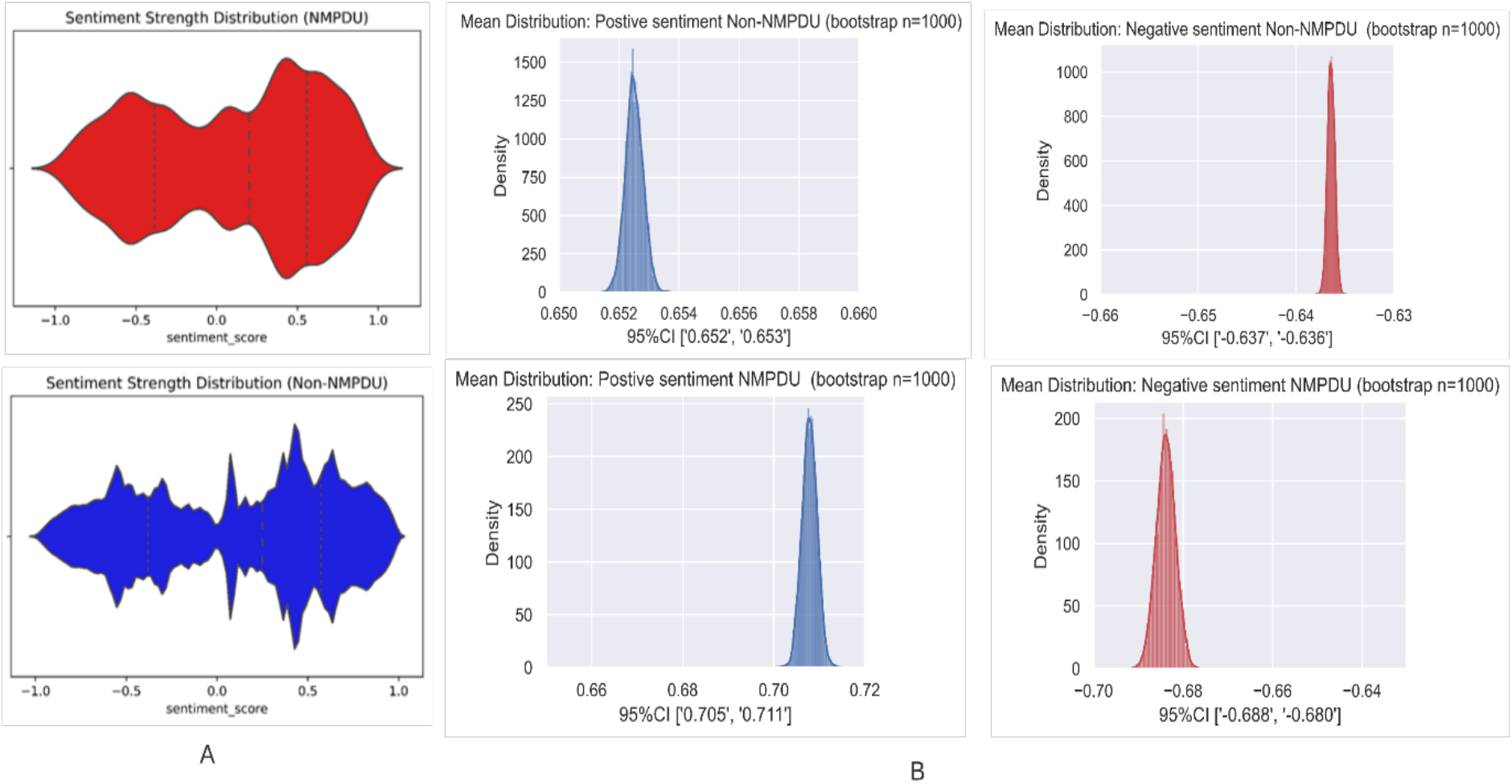
A: Sentiment strength distributions for NMPDU and non-NMPDU posts, B: Comparison of the means and confidence intervals of extreme positive tweets (sentiment score >0.5) and negative tweets (sentiment score <− -0.5) in both Non-NMPDU and NMPDU categories.

### Personal and social concern analysis

We measured differences between the tweets from the NMPDU and non-NMPDU groups in terms of the following content dimensions: personal concern (e.g., work, leisure, home, money, religion, and death), social content (e.g., family and friends), time orientation content (e.g., past focus), core drive content (e.g., achievement), and biological process content (e.g., health and body). Table 4 presents the medians, Mann– Whitney U test results, and the effect sizes of the comparisons between the NMPDU group and the control group tweets. In addition, we present comparisons between the groups for each medication category (i.e., opioids, benzodiazepines, stimulants, and polysubstance) in Supplementary S.4.

**Table 4.** Comparison of the personal and social concern content between the users from the NMPDU and control groups

| Content | Variables | Median: NMPDU group | Median: Control group | Different (NMPDU versus control) Mann–Whitney U test results |  |
| --- | --- | --- | --- | --- | --- |
|  |  |  |  | P-value (p) | Effect sizes (r) |
| **Social content** | Family | 0.0333 | 0.025 | <0.001 | 0.19* |
|  | Friend | 0.0187 | 0.020 | >0.001 | - |
| **Time orientation content** | Past focus | 0.241 | 0.140 | <0.001 | 0.59*** |
| **Core drive content** | Achieve | 0.098 | 0.204 | <0.001 | 0.63*** |
| **Biological process content** | Health | 0.047 | 0.056 | <0.001 | 0.15* |
|  | Body | 0.112 | 0.086 | <0.001 | 0.28* |
| **Personal concern content** | Work | 0.098 | 0.215 | <0.001 | 0.60*** |
|  | Leisure | 0.120 | 0.272 | <0.001 | 0.71**** |
|  | Home | 0.031 | 0.058 | <0.001 | 0.50*** |
|  | Money | 0.055 | 0.086 | <0.001 | 0.41** |
|  | Religion | 0.032 | 0.041 | <0.001 | 0.23* |
|  | Death | 0.018 | 0.016 | >0.001 | - |

Size effects(r): \*Small difference ( $0.1 \leq r < 0.3$ ). \*\*Medium difference ( $0.3 \leq r < 0.5$ ). \*\*\*Large difference ( $0.5 \leq r < 0.7$ ). \*\*\*\*Very large difference ( $r \geq 0.7$ )<sup>36</sup>

The users from the NMPDU group express significantly more social content related to family (p < 0.001, r = 0.19) than the users from the non-NMPDU group, but no significant difference is observed between the groups in content related to friends (p > 0.001) (Table 4). The comparison in the personal concern content demonstrate that the users from NMPDU group express significantly less personal concern content related to work (p < 0.001, r = 0.60), leisure (p < 0.001, r = 0.71), home (p < 0.001, r = 0.50), money (p < 0.001, r = 0.41), and religion (p < 0.001, r = 0.23) compared to the users from the control group. No significant difference is found in the death variable (p > 0.001). For biological process content, the users from the NMPDU group tend to use less content related to health (p < 0.001, r = 0.15) and use more content related to the body (p < 0.001, r = 0.28) than the users from the control group. Comparing both groups based on time orientation content shows that the users from the NMPDU group tend to discuss significantly more content related to the past (p < 0.001, r = 0.59) than the users from the control group. Finally, the users from the NMPDU group express significantly less core drive content related to achievement (p < 0.001, r = 0.63) compared to the users from the control group.

### Gender differences in concerns within the NMPDU group

Female within the NMPDU express significantly more social content related to family (p < 0.001, r = 0.27) and friends (p > 0.001, r = 0.50) compared to the male NMPDU users (Table 5, Figure 2). Also, no significant gender differences are observed in personal concern content related to all but the home variable, with female NMPDU users expressing more content related to home (p > 0.001, r = 0.41) compared to the male NMPDU users. For biological process content, the female NMPDU users tend to use more content related to health (p < 0.001, r = 0.37), while no significant gender difference exists in content related to the body. For time orientation content, the female NMPDU users tend to discuss significantly more content related to the past (p < 0.001, r = 0.21) than the male NMPDU users. Finally, there is no significant gender difference in core drive content related to achievement.

**Table 5.** Comparison of the personal and social concern content between the male and female users from the NMPDU group

| Content | Variables | Median: Male NMPDU group | Median: Female NMPDU group | Different (Male NMPDU versus Female NMPDU I) Mann–Whitney U test results |  |
| --- | --- | --- | --- | --- | --- |
|  |  |  |  | P-value (p) | Effect sizes (r) |
| Social content | Family | 0.029 | 0.038 | <0.001 | 0.27* |
|  | Friend | 0.013 | 0.026 | <0.001 | 0.50*** |
| Time orientation content | Past focus | 0.229 | 0.267 | <0.001 | 0.21* |
| Core drive content | Achieve | 0.105 | 0.114 | >0.001 | - |
| Biological process content | Health | 0.041 | 0.057 | <0.001 | 0.37** |
|  | Body | 0.108 | 0.120 | >0.001 | - |
| Personal concern content | Work | 0.092 | 0.110 | >0.001 | - |
|  | Leisure | 0.121 | 0.125 | >0.001 | - |
|  | Home | 0.025 | 0.040 | <0.001 | 0.41** |
|  | Money | 0.056 | 0.056 | >0.001 | - |
|  | Religion | 0.033 | 0.033 | >0.001 | - |
|  | Death | 0.018 | 0.018 | >0.001 | - |

Size effects(r): \*Small difference ( $0.1 \leq r < 0.3$ ). \*\*Medium difference ( $0.3 \leq r < 0.5$ ). \*\*\*Large difference ( $0.5 \leq r < 0.7$ ).
\*\*\*\*Very large difference ( $r \geq 0.7$ )<sup>36</sup>

**Figure 2:**
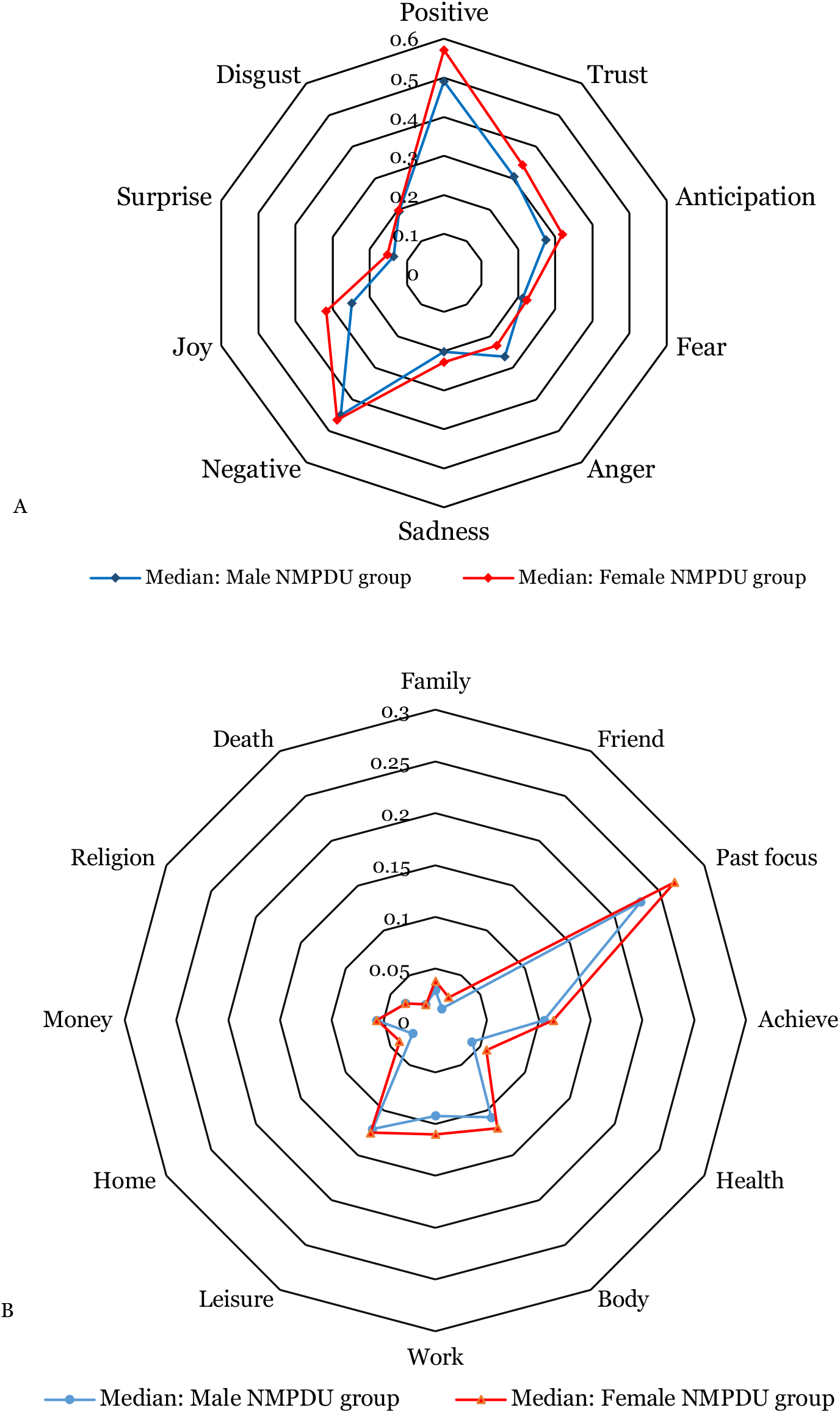
Summary comparison between the male and female users from the NMPDU group. (A) Emotional dimensions, (B) the personal and social concern content dimensions

### Potential reasons for NMPDU

Table 6 shows the summary of the potential reasons for NMPDU and frequently used keywords indicating these reasons for each medication category. We applied latent Dirichlet allocation (LDA), a topic modeling method, for each category separately. We then examined the word clusters in each set of the sub-topics obtained via LDA (see Supplementary S.5). Subsequently, we interpreted the identified topics and selected potential reasons. These reasons were inferred by manually inspecting the frequent words within each category qualitatively, guided by the National Survey on Drug Use and Health (NSDUH) surveys^37^.

**Table 6.**
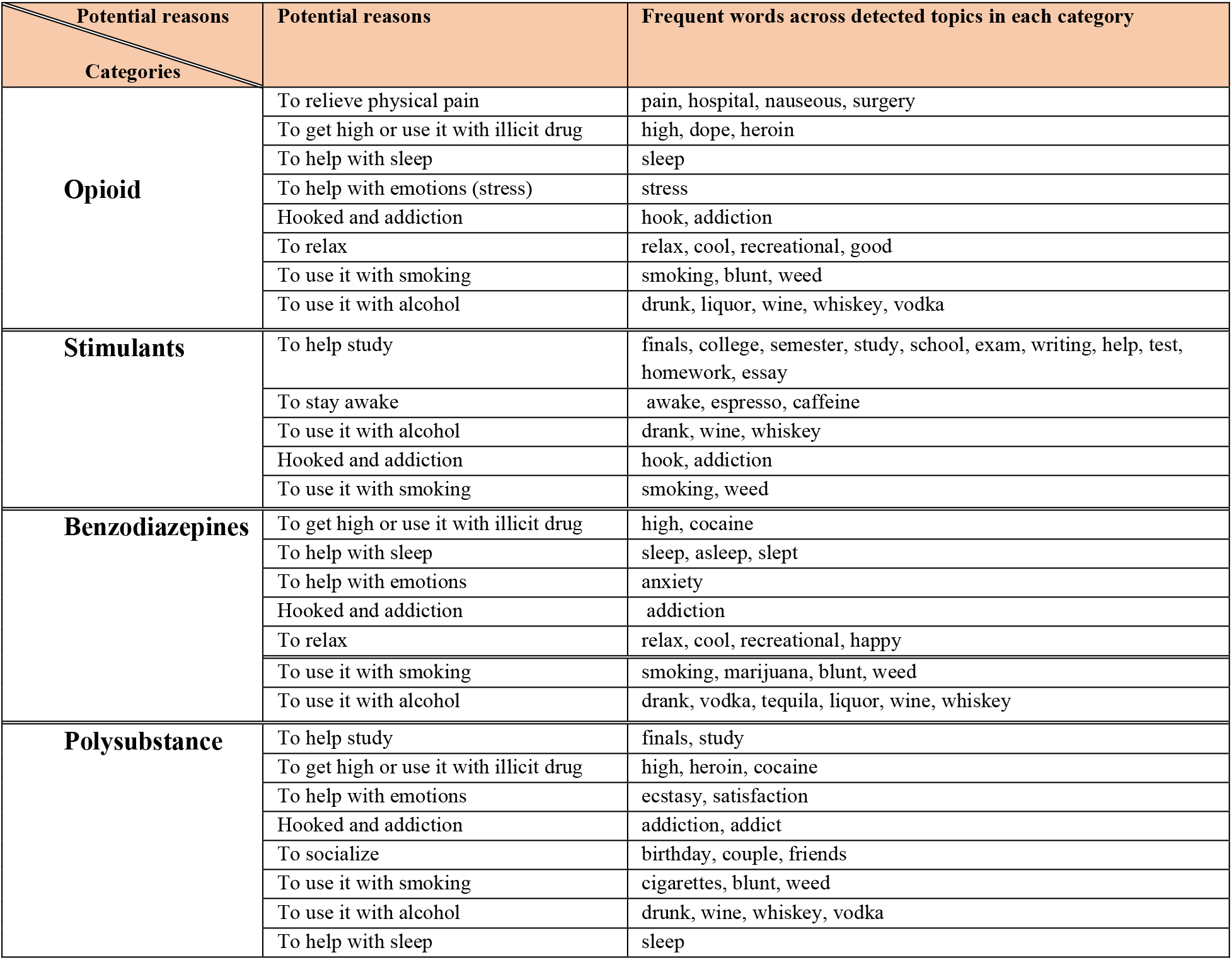
Potential reasons for NMPDU extracted from topic modeling content analysis

## Discussions

Our emotion analysis showed significant differences in the emotion-indicating expressions in the tweets between users from the NMPDU and control groups. Relative to users from the non-NMPDU group, users from the NMPDU group posted more emotionally negative content and less emotionally positive content in their Twitter posts. Relative to the non-NMPDU tweets, the NMPDU tweets contained higher numbers of extremely polarized (positive or negative) tweets, indicating possible emotional triggers associated with NMPDU. We also found significant differences in the contents shared between female and male nonmedical users of prescription drugs. Compared to the female users, the male users expressed higher anger and lower positivity, joy, anticipation, and sadness in their posted contents. In terms of social and personal content, compared to the male users, the female users shared more content related to social life (friends and family), health, and personal concern (home). Interestingly, while there were unique and detectable differences in the contents between male and female nonmedical prescription drug users, the differences were consistent across different drug categories. These findings perhaps indicate that the underlying reasons behind NMPDU may be associated with cohort-level behavioral characteristics more than the properties of the substances themselves. From the perspective of public health, the insights obtained through this large-scale analysis of social media data may help customize awareness and intervention programs to targeted cohorts in order to mitigate the population-level impacts of NMPDU.

Our study adds to the growing body of literature focusing on the intersection of substance use and behavioral health. The findings from our large-scale social media analyses are consistent with previous results from a survey-based study^38^ that showed that those who reported specific feelings, such as hopelessness, sadness, or depression, are more likely to report nonmedical use of opioids stimulants, sedatives, and antidepressants. The consistency in findings across studies demonstrates the utility of social media for NMPDU surveillance—in this case, surveillance may not only help estimate NMPDU at the population level but also provide in-depth insights into the emotional and behavioral drivers of NMPDU. Social media-based surveillance systems have the potential of operating in close to real-time while costing less than traditional surveillance systems and have the ability to include hard-to-reach populations (e.g., people without health coverage in the US). While social media-based surveillance systems will not replace the traditional ones, they may offer complimentary yet important information.

A previous study reported an association between the uses of emotional words (user-generated natural language) and individuals’ experiences (individual differences in mood, personality, and physical and emotional well-being)^26^. The study showed that negative emotion words were associated with psychological distress and poor physical health, whereas higher positive emotion words are associated with better well-being and physical health. Thus, although our study did not directly examine such an association among the NMPDU users on Twitter, we posit that the higher numbers of negative emotion words of the users from the NMPDU group are likely associated with greater psychological distress and poorer physical health compared to their non-NMPDU counterparts, a hypothesis that we plan to study in future work.

Our study also demonstrates that potential specific reasons behind NMPDU may be derived from social media data, and this finding may have major public health implications. This information can be useful to policymakers for implementing measures for drug use prevention, intervention, and treatment in their communities^37^. As shown in Table 6 and elaborated in Supplementary material S.5, “*to relieve pain*” is one reason for the NMPDU of opioids, indicating that opioids are often used for treating pain and that not all prescription opioid use is for recreational reasons or due to addiction. The reason “*to help with emotions*” is common in the NMPDU of opioids and benzodiazepines, suggesting that these two categories of medications are potentially used nonmedically for coping with emotional anguish. “*To help with sleep*” is reported as a reason for the NMPDU of opioids, benzodiazepines, and polysubstance, suggesting that many people nonmedically use these substances for addressing their sleep problems. Over the recent years, the co-ingestion of opioids with benzodiazepines has led to rising overdose-related deaths^39^. Since our findings indicate that many people may be using these substances for addressing sleep problems, more efforts are called for to educate the general public about non-pharmacological, safer strategies to mitigate sleep problems/improve sleep quality. Healthcare providers could help identify and intervene with the root causes of their patients’ sleep problems. These efforts could contribute to reducing drug overdose-related mortality. The topic analysis also suggests the nonmedical use of stimulants are often to enhance educational performance and for staying awake. Past research has shown that nonmedical use of prescription stimulants, such as Adderall®, is widespread among college students^40,41^, and our findings agree with these studies. Overdose deaths due to stimulants (prescription and illicit, particularly co-use with fentanyl and other opioids) are rapidly increasing in the US, which might be partly attributed to the many years of widespread prescription stimulant use in educational settings^42^. Students could benefit from awareness programs in educational institutions or adolescent/young adult healthcare settings to prevent adverse, often fatal, health consequences caused by stimulant use. The topics associated with all the medication categories are indicative of co-use of prescription drugs with other legal substances such as alcohol and tobacco, and indicative of NMPDU due to substance use disorder. Specifically for opioids, benzodiazepines and polysubstances, there are topics that are indicative of co-use with illicit substances such as cocaine and heroin. Topics associated with nonmedical use of benzodiazepines are indicative of their use for relieving stress. Finally, topics associated with polysubstances are indicative of their use in social settings.

As mentioned earlier, social media provides a unique opportunity to study NMPDU at a macro level in close to real-time. Although social media data presents its own challenges, such as the use of colloquial expressions and non-standard spelling variants, advances in machine learning and NLP methods have enabled us to leverage the vast knowledge encapsulated in this resource for public health surveillance. The drug overdose death rates published by the CDC every year include a close to two-year ‘lag’, meaning that currently in 2021, we only have complete data up to 2020. Public health measures designed based on such laggy information may not be as effective as close-to-real-time measures, and this is one area where social media mining may contribute. There is also the potential to integrate social media data with traditional data sources (e.g., survey data) to obtain a complete picture of population-level substance use.

Our study has several limitations. A major limitation is that data from social media may not be well representative of the overall population. Social media users tend to be younger and technologically savvy, resulting in a biased sample. However, it is also unlikely that any other resource matches the scale and reach of social media, and as the demographics shift, more and more older adults are reachable via social media^43^. As mentioned above, the triangulation of social media and traditional survey data (or any other offline data source) to study NMPDU can help minimize the potential biases in the representative samples. There are also limitations associated with the methods we employed. We applied topic modeling to discover potential reasons for NMPDU. Unlike supervised methods (e.g., classification), which can be evaluated against human experts, it is not possible to thoroughly evaluate the performance of topic modeling. The performance of topic modeling may vary, and there is no mechanism to evaluate such approaches in a task-oriented manner. Also, our study findings are dependent on the classification performances of the machine-learning and NLP pipelines. The performances of these methods are not 100% accurate and may add further biases in the downstream analyses.

## Methods

### Data collection

For NMPDU, similar to our previous study^44^, which discussed designing a pipeline tool to collect NMPDU data from social media, we used a list of keywords (see Supplementary S.2) after consultation with the toxicology expert of our study (JP). We included a list of prescription drugs, including opioids, benzodiazepines, and central nervous system stimulants, which are known for their misuse/abuse potential. We also included polysubstance users (users who have self-reported using multiple drugs at the same time). First, we extracted approximately 3,287,703 tweets that contained a list of identified keywords related to prescription drugs from March 6, 2018, to January 14, 2020, to be the seeds for the collection of NMPDU users. We used an advanced NLP-based model (see NMPDU classification model) to classify the tweets automatically into one of four categories: NMPDU, consumption, mention, and unrelated. Mining NMPDU information from social media is more challenging than mining illicit drug use information, particularly because consumption of prescription drugs does not automatically indicate abuse. We extracted the complete publicly available user profiles (i.e., all publicly available tweets) of users who posted the NMPDU tweets to build our experimental group (NMPDU users). We removed any user with less than 500 tweets. As shown in Table 1, we collected 49,833 NMPDU users with approximately 82 million tweets. For non-NMPDU users (control group), we randomly extracted publicly available profiles whose gender was identified via Liu & Ruths^45^ and *Volkova et al*. ^46^. and who have not mentioned any identified prescription drug keywords in their profiles, resulting in 37,885 non-NMPDU users with approximately 55 million tweets. Overall, we included complete publicly available profiles of 87,718 users with approximately 137 million tweets.

### NMPDU classification model

We used an NLP text classification model developed and validated in our previous research ^47^ to distinguish NMPDU from non-NMPDU tweets. The model uses RoBERTa—a transformer-based language model—to classify tweets into 1) NMPDU (potential misuse), 2) consumption (consumption but no evidence of nonmedical use), 3) mention (drug mentioned but no evidence of consumption), and 4) unrelated. Overall, the NMPDU classification model has an accuracy of 82.32%, and the F_1_ score for each class are NMPDU 65%, consumption 91%, mention 88%, and unrelated 90%).

### Gender Label

The gender of the non-NMPDU users (control group) was released publicly on Twitter and identified via previous work (Liu & Ruths^45^ and Volkova et al. ^46^). The gender of the NMPDU users was inferred using an NLP text classification model developed in the authors’ previous work ^48^. This model uses users’ metadata (name, screen name, and description) and tweets to label the users using a binary gender paradigm (i.e., male and female) and has an accuracy of 94.4% on NMPDU users.

### Emotion analysis

For emotion analysis, we used the word emotion lexicon curated by the NRC^32^. The lexicon is a list of approximately 14,000 English words and their associations with eight basic emotions (anger, fear, anticipation, trust, surprise, sadness, joy, and disgust) according to Plutchik’s research on basic emotions^49^. The annotations are manually done by crowdsourcing^32^. The emotion lexicon has been used to study and categorize the emotion in the Twitter text by several prior studies^50,51,52^, and it is considered the benchmark for this domain of data.

### Sentiment analyses of NMPDU tweets (sentiment score classifier)

We used VADER^53^, an open-source Twitter sentiment model, which assigns numerical sentiment scores between +1 (extremely positive sentiment) and −1 (extremely negative sentiment) to each tweet. VADER has been used as the sentiment analyzer in several previous studies^54,55^. Furthermore, a survey research^56^ that compared the results of two classes of sentiment classifiers on four datasets from Twitter concluded that VADER has the best performance, with an overall accuracy of 99.04% (positive class: precision = 99.16%, recall = 99.16%, F1 scores = 99.31%; negative class: precision = 98.77%, recall = 98.12%, F1 scores = 98.88%).

### Personal and social concern analysis

We used the validated Linguistic Inquiry and Word Count (LIWC), which characterizes words in psychologically meaningful categories^57^. LIWC is used to analyze several varieties of text, including social media text. The LIWC lexicon, which is designed to measure several behavioral and psychological dimensions from text, has been used in several prior studies ^58,59,50^ for physiological measures of well-being analysis from social media.

### Topic Modeling

For topic modeling, we utilize LDA ^60^, an effective unsupervised method that assumes that each document in a large dataset comprises sub-topics represented by the words they contain. We initially cleaned the tweets by removing hyperlinks, digits, and stop words. Then, to decide the ideal number of topics for our model, we executed multiple models with different hyperparameters (number of topics = 5, 15, 20, 30, 40, and 50). We then inspected the word clusters in each set of sub-topics and determined the most salient set of topics. Subsequently, we selected the 20-topic model. By using the frequent words in each sub-topic, we qualitatively estimated the potential reasons for nonmedical uses for each category of NMPDU (see Supplementary S.5).

## Data Availability

Aggregated data and code for the experiments are available. Details are provided in the Data Availability and Code Availability statements within the mauscript.

## Data availability

The data used in this study are publicly available from Twitter. However, it cannot be distributed by the authors. Statistical data extracted from the Twitter content reported in this paper’s findings and the source code needed to replicate the findings can be downloaded from the following <u>code link</u>. The authors can provide the researchers with the IDs required for downloading tweets directly from the Twitter application programming interface upon reasonable request.

## Code availability

To aid reproducible research, the code and aggregated data of this study are freely available from <u>code link</u>. Additional data and information are available from the authors upon reasonable request.

## Acknowledgements

Research reported in this publication is supported by the National Institute on Drug Abuse (NIDA) of the National Institutes of Health (NIH) under award number R01DA046619. The content is solely the responsibility of the authors and does not necessarily represent the official views of the NIH.

## Authors’ contributions

MAA, YCY and AS designed the experiments. MAA, YCY, YG, and AS conducted the data collection, analysis, and evaluations. MAA and AS conducted manuscript writing. JP, SK, JSL provided medical domain expertise, helped interpret relevant findings, discussed and analyzed the results.

## Competing interests

The authors declare no competing interests.

## Notes

### Competing Interest Statement

The authors have declared no competing interest.

### Author Declarations

Emory University Institutional Review Board (exempt category 4)

