## Supplementary for "Large-scale social media language analysis reveals emotions and behaviours associated with nonmedical prescription drug use"

^1^Department of Biomedical Informatics, School of Medicine, Emory University, Atlanta, GA, United States. ^2^Computer Science Emory University Atlanta GA, United States. School of Nursing, Emory University, Atlanta, GA, United States. ^4^Department of Emergency Medicine, School of Medicine, Oregon Health & Science University, Portland, OR, United States; ^5^Department of Emergency Medicine, Perelman School of Medicine, University of Pennsylvania, Philadelphia, PA, United States. ^6^Department of Biomedical Engineering, Georgia Institute of Technology and Emory University, Atlanta, GA, United States.

### **S.1 Definitions and Terminology**

In literature, the terms “nonmedical use,” “abuse,” and “misuse” are often used interchangeably to describe the nonmedical use of prescription drugs (i.e., prescription drug abuse or prescription drug misuse) [1–5]. According to the National Survey on Drug Use and Health, the nonmedical use/misuse/abuse of prescription drugs can refer to the use of prescription drug that a doctor did not direct the respondent to use, including the consumption of dosage other than that prescribed or the consumption with other prescription drugs [6]. The full discussion of the guideline for annotating and creating high-quality social media corpora, which was used to train the NMPDU classifier, is provided in our previous study [1].

*NMPDU users (Experimental Group)*: We define NMPDU users as users who have posted at least one tweet that is labeled an NMDPU tweet, including prescription medication (PM)-identified keywords, during the data collection process (March 6, 2018–January 14, 2020) and users who have more than 500 tweets in their Twitter profile. The complete available Twitter profile is included in the analysis only if the NMPDU tweet is still available at the time of profile extraction.

*NMPDU users (Control Group):* We selected random tweets (without any selection criteria) and from them extracted the complete profile of users who posted the tweets. The final inclusion criteria were the users who did not mention PM-identified keywords in their profile, and the users had more than 500 tweets in their profile.

### **S.2 Data collection**

#### **S.2.1 Keywords list**

The identified keywords used for extracting tweets (generic and brand names) [1] were as follows:

**Opioids**

| Generic name | Brand Name(s) |
| --- | --- |
| Oxycodone | Oxycontin, Percocet |
| Methadone | Dolophine |
| Morphine | Avinza |
| Tramadol | Conzip |
| Hydrocodone | Vicodin, Zohydro |
| Buprenorphine/naloxone | Suboxone |

**Stimulants**

| Generic name | Brand Name(s) |
| --- | --- |
| Amphetamine mixed salts | Adderall |
| Lisdexamfetamine | Vyvanse |
| Methylphenidate | Ritalin |

**Benzodiazepines**

| Generic name | Brand Name(s) |
| --- | --- |
| Diazepam | Valium |
| Alprazolam | Xanax |
| Clonazepam | Klonopin |
| Lorazepam | Ativan |

**Polysubstance:** Tweets that contain drugs from different categories.

### **S.3 Normality and statistical tests**

We used histogram as an initial graphical test to check if data samples are normally distributed. Histogram is a commonly used test for carrying out a quick check on the normality of data distribution. Data are said to be normally distributed if they have a Gaussian-like shape (bell shape). The assessment is done qualitatively, but in many cases, deciding if the data are normally distributed or not is difficult. Therefore, we used Anderson–Darling (AD) as the statistical test for normality check.

The AD test is used to check if the population from which the data samples are drawn is normally distributed [7] [8]. It adds an adjustment to the Kolmogorov–Smirnov (K–S) test to provide more weight to the tails. The one-sample AD test statistic is non-directional and calculated using the following formula [8]:

H0: The population from which the data samples are drawn is normally distributed.

Ha: The population from which the data samples are drawn is not normally distributed.

$$AD^{2}=-N-S$$

$$S=\sum_{i=1}^{N} \frac{2i-1}{N}\left( \log\left( X_{i} \right)+\log\left( 1-X_{n-i+1} \right) \right)$$

Note that S is

Moreover, $x_{n}$is the ordered elements of the sample, which has a size of$n$. $F(x)$is the [cumulative distribution function](https://www.itl.nist.gov/div898/handbook/eda/section3/eda362.htm#CDF) that is used for comparison with the sample. The null hypothesis is rejected if $AD$ is larger than the critical value $AD\alpha$ at a given $\alpha$(critical values for different sample sizes can be obtained [9]) [8].

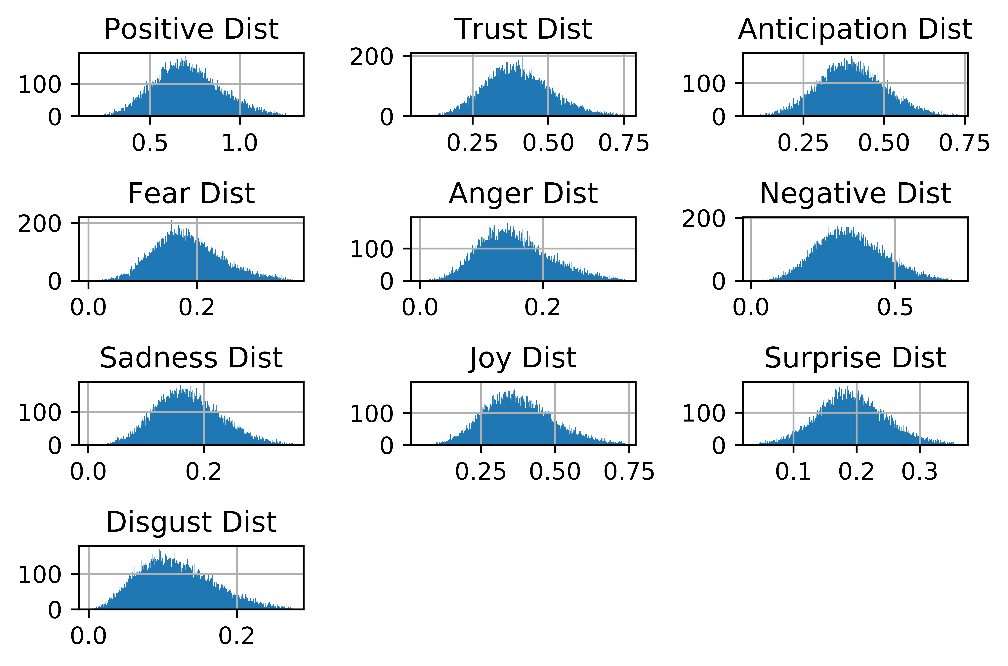

**Figure 1 Histogram distribution (Dist) for the emotion control group**

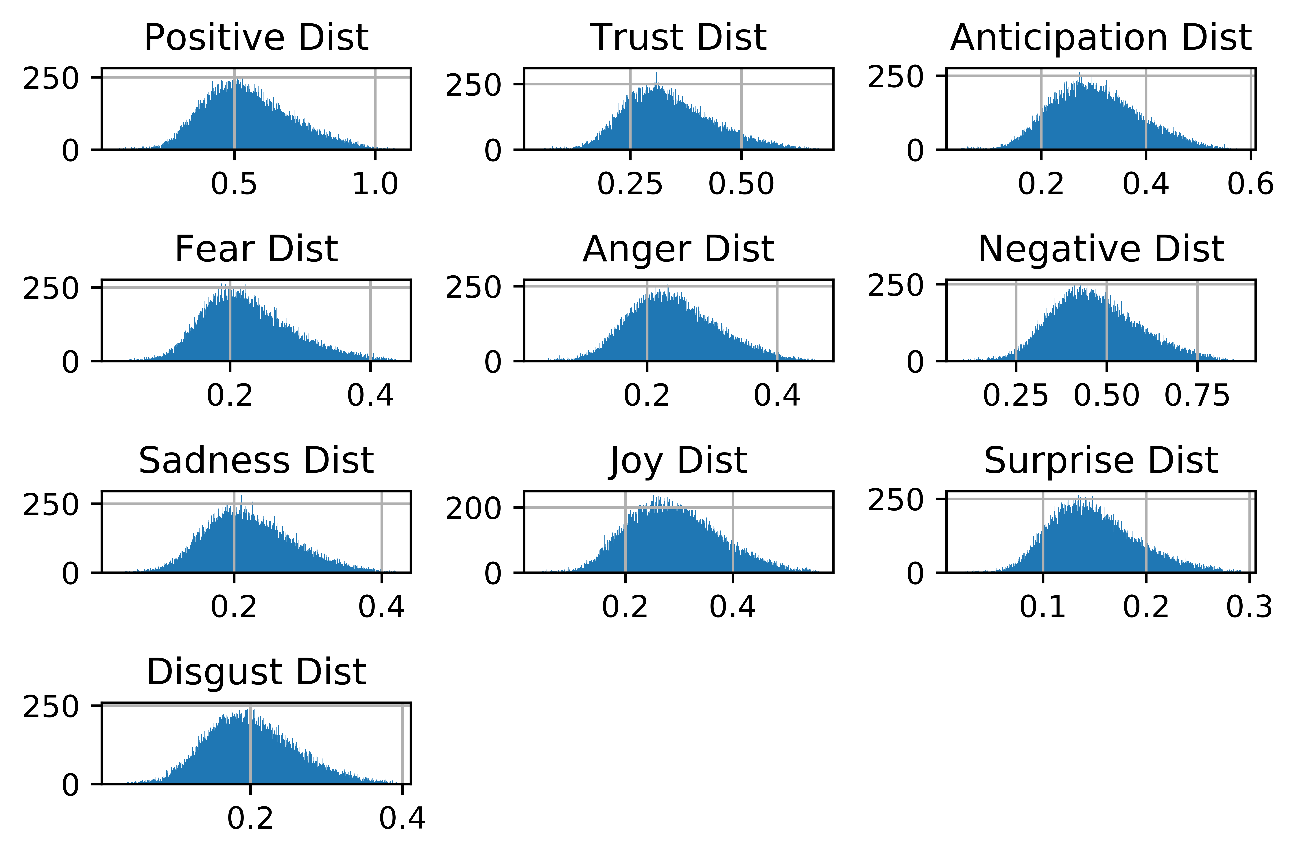

**Figure 2 Histogram distribution (Dist) for the emotion NMPDU group**

**Table 2 Normality test (AD) for the NMPDU Group and Control Group (emotion variables)**

| **Emotional variables** | **NMPDU Group** | **Control Group (Non-NMPDU Group)** |
| --- | --- | --- |
| **Positive** | Statistic: 51.74* > greater than the critical values:-  15% (0.576,), 10% (0.656), 5% (0.787), 2.5(0.918)%, 1 (1.091)%  *Data is not normally distributed (reject H0) | Statistic: 41.79* > greater than the critical values:-  *15% (0.576,), 10% (0.656), 5% (0.787), 2.5(0.787)%, 1 (1.092)%*  *Data is not normally distributed (reject H0) |
| **Trust** | Statistic: 60.39* > greater than the critical values:-  15% (0.576,), 10% (0.656), 5% (0.787), 2.5(0.918)%, 1 (1.091)%  *Data is not normally distributed (reject H0) | Statistic: 61.57* > greater than the critical values:-  *15% (0.576,), 10% (0.656), 5% (0.787), 2.5(0.787)%, 1 (1.092)%*  *Data is not normally distributed (reject H0) |
| **Anticipation** | Statistic: 38.87* > greater than the critical values:-  15% (0.576,), 10% (0.656), 5% (0.787), 2.5(0.918)%, 1 (1.091)%  *Data is not normally distributed (reject H0) | Statistic: 17.58* > greater than the critical values:-  *15% (0.576,), 10% (0.656), 5% (0.787), 2.5(0.787)%, 1 (1.091)%*  *Data is not normally distributed (reject H0) |
| **Fear** | Statistic: 38.90* > greater than the critical values:-  15% (0.576,), 10% (0.656), 5% (0.787), 2.5(0.918)%, 1 (1.091)%  *Data is not normally distributed (reject H0) | Statistic: 97.97* > greater than the critical values:-  *15% (0.576,), 10% (0.656), 5% (0.787), 2.5(0.787)%, 1 (1.092)%*  *Data is not normally distributed (reject H0) |
| **Anger** | Statistic: 11.22* > greater than the critical values:-  15% (0.576,), 10% (0.656), 5% (0.787), 2.5(0.918)%, 1 (1.091)%  *Data is not normally distributed (reject H0) | Statistic: 130.411* > greater than the critical values:-  *15% (0.576,), 10% (0.656), 5% (0.787), 2.5(0.787)%, 1 (1.092)%*  *Data is not normally distributed (reject H0) |
| **Negative** | Statistic: 11.20* > greater than the critical values:-  15% (0.576,), 10% (0.656), 5% (0.787), 2.5(0.918)%, 1 (1.091)%  *Data is not normally distributed (reject H0) | Statistic: 57.94* > greater than the critical values:-  *15% (0.576,), 10% (0.656), 5% (0.787), 2.5(0.787)%, 1 (1.092)%*  *Data is not normally distributed (reject H0) |
| **Sadness** | Statistic: 22.95* > greater than the critical values:-  15% (0.576,), 10% (0.656), 5% (0.787), 2.5(0.918)%, 1 (1.091)%  *Data is not normally distributed (reject H0) | Statistic: 50.28* > greater than the critical values:-  *15% (0.576,), 10% (0.656), 5% (0.787), 2.5(0.787)%, 1 (1.092)%*  *Data is not normally distributed (reject H0) |
| **Joy** | Statistic: 41.27* > greater than the critical values:-  15% (0.576,), 10% (0.656), 5% (0.787), 2.5(0.918)%, 1 (1.091)%  *Data is not normally distributed (reject H0) | Statistic: 80.46* > greater than the critical values:-  *15% (0.576,), 10% (0.656), 5% (0.787), 2.5(0.787)%, 1 (1.092)%*  *Data is not normally distributed (reject H0) |
| **Surprise** | Statistic: 48.30* > greater than the critical values:-  15% (0.576,), 10% (0.656), 5% (0.787), 2.5(0.918)%, 1 (1.091)%  *Data is not normally distributed (reject H0) | Statistic: 14.17* > greater than the critical values:-  *15% (0.576,), 10% (0.656), 5% (0.787), 2.5(0.787)%, 1 (1.092)%*  *Data is not normally distributed (reject H0) |
| **Disgust** | Statistic: 9.84* > greater than the critical values:-  15% (0.576,), 10% (0.656), 5% (0.787), 2.5(0.918)%, 1 (1.091)%  *Data is not normally distributed (reject H0) | Statistic: 141.92* > greater than the critical values:-  *15% (0.576,), 10% (0.656), 5% (0.787), 2.5(0.787)%, 1 (1.092)%*  *Data is not normally distributed (reject H0) |

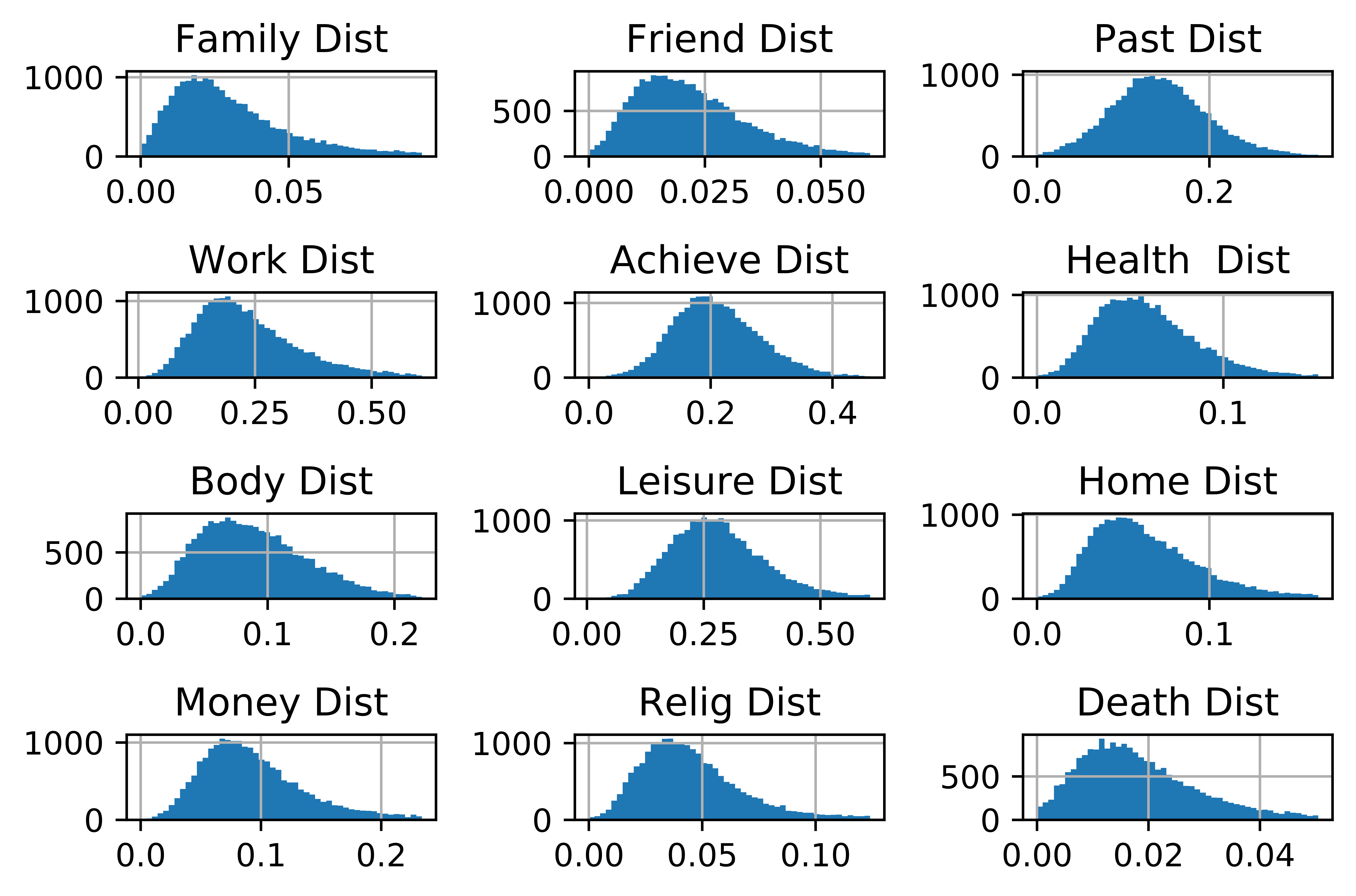

**Figure 3 Histogram distribution (Dist) of the control group (personal and social variables)**

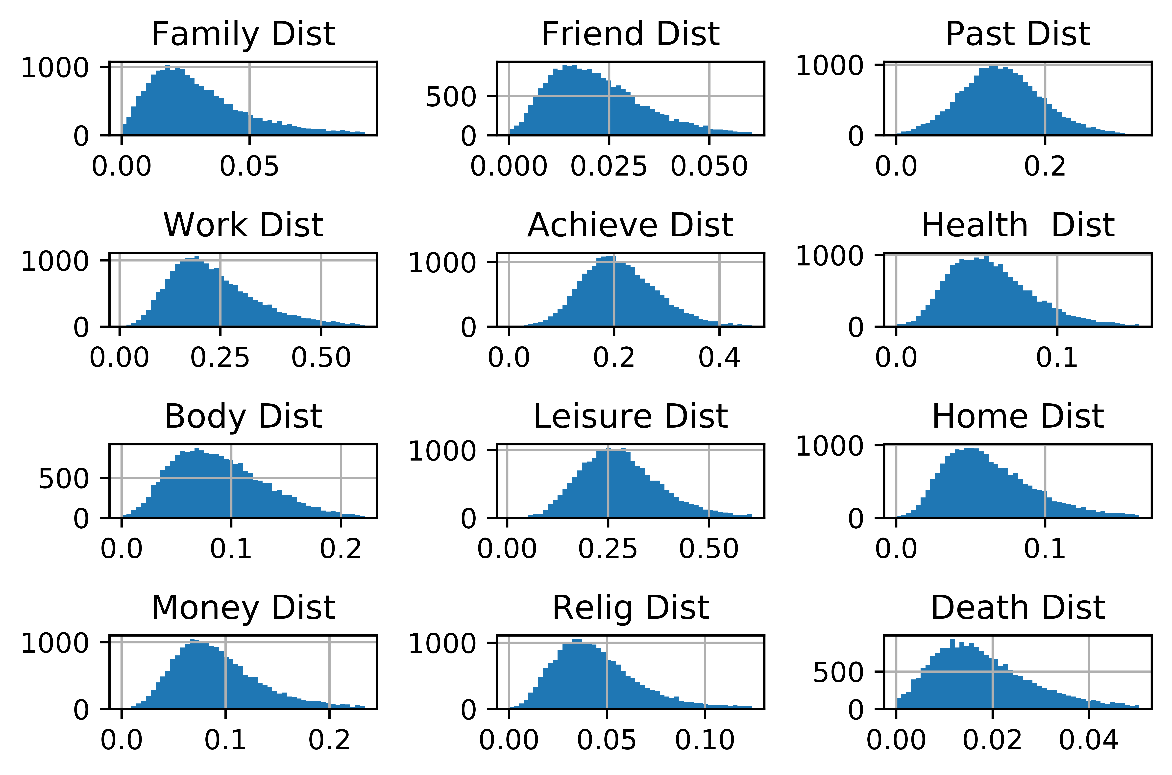

**Supplementary Figure 4 Histogram distribution (Dist) for social and personal variables of the NMPDU group**

**Table 1: Normality test (AD) for the NMPDU Group and Control Group (personal and social variables)**

| **personal and social variables** | **NMPDU Group** | **Control Group (Non-NMPDU Group)** |
| --- | --- | --- |
| **Family** | Statistic: 58.63* > greater than the critical values:-  15% (0.576,), 10% (0.655), 5% (0.786), 2.5(0.9187)%, 1 (1.091)%  *Data is not normally distributed (reject H0) | Statistic: 396.9* > greater than the critical values:-  15% (0.576,), 10% (0.656), 5% (0.787), 2.5(0.787)%, 1 (1.092)%  *Data is not normally distributed (reject H0) |
| **Friend** | Statistic: 111.80* > greater than the critical values:-  15% (0.576,), 10% (0.655), 5% (0.786), 2.5(0.9187)%, 1 (1.091)%  *Data is not normally distributed (reject H0) | Statistic: 193.2* > greater than the critical values:-  15% (0.576,), 10% (0.656), 5% (0.787), 2.5(0.787)%, 1 (1.092)%  *Data is not normally distributed (reject H0) |
| **Past** | Statistic: 12.53* > greater than the critical values:-  15% (0.576,), 10% (0.655), 5% (0.786), 2.5(0.9187)%, 1 (1.091)%  *Data is not normally distributed (reject H0) | Statistic: 22.5* > greater than the critical values:-  15% (0.576,), 10% (0.656), 5% (0.787), 2.5(0.787)%, 1 (1.091)%  *Data is not normally distributed (reject H0) |
| **Work** | Statistic: 133.98* > greater than the critical values:-  15% (0.576,), 10% (0.655), 5% (0.786), 2.5(0.9187)%, 1 (1.091)%  *Data is not normally distributed (reject H0) | Statistic: 246.67* > greater than the critical values:-  15% (0.576,), 10% (0.656), 5% (0.787), 2.5(0.787)%, 1 (1.092)%  *Data is not normally distributed (reject H0) |
| **Achieve** | Statistic: 59.09* > greater than the critical values:-  15% (0.576,), 10% (0.655), 5% (0.786), 2.5(0.9187)%, 1 (1.091)%  *Data is not normally distributed (reject H0) | Statistic: 52.95* > greater than the critical values:-  15% (0.576,), 10% (0.656), 5% (0.787), 2.5(0.787)%, 1 (1.092)%  *Data is not normally distributed (reject H0) |
| **Health** | Statistic: 54.37* > greater than the critical values:-  15% (0.576,), 10% (0.655), 5% (0.786), 2.5(0.9187)%, 1 (1.091)%  *Data is not normally distributed (reject H0) | Statistic: 136.72* > greater than the critical values:-  15% (0.576,), 10% (0.656), 5% (0.787), 2.5(0.787)%, 1 (1.092)%  *Data is not normally distributed (reject H0) |
| **Body** | Statistic: 6.90* > greater than the critical values:-  15% (0.576,), 10% (0.655), 5% (0.786), 2.5(0.9187)%, 1 (1.091)%  *Data is not normally distributed (reject H0) | Statistic: 118.17* > greater than the critical values:-  15% (0.576,), 10% (0.656), 5% (0.787), 2.5(0.787)%, 1 (1.092)%  *Data is not normally distributed (reject H0) |
| **Leisure** | Statistic: 72.02* > greater than the critical values:-  15% (0.576,), 10% (0.655), 5% (0.786), 2.5(0.9187)%, 1 (1.091)%  *Data is not normally distributed (reject H0) | Statistic: 80.47* > greater than the critical values:-  15% (0.576,), 10% (0.656), 5% (0.787), 2.5(0.787)%, 1 (1.092)%  *Data is not normally distributed (reject H0) |
| **Home** | Statistic: 91.89* > greater than the critical values:-  15% (0.576,), 10% (0.655), 5% (0.786), 2.5(0.9187)%, 1 (1.091)%  *Data is not normally distributed (reject H0) | Statistic: 214.82* > greater than the critical values:-  15% (0.576,), 10% (0.656), 5% (0.787), 2.5(0.787)%, 1 (1.092)%  *Data is not normally distributed (reject H0) |
| **Money** | Statistic: 54.04* > greater than the critical values:-  15% (0.576,), 10% (0.655), 5% (0.786), 2.5(0.9187)%, 1 (1.091)%  *Data is not normally distributed (reject H0) | Statistic: 227.84* > greater than the critical values:-  15% (0.576,), 10% (0.656), 5% (0.787), 2.5(0.787)%, 1 (1.092)%  *Data is not normally distributed (reject H0) |
| **Religion** | Statistic: 35.45* > greater than the critical values:-  15% (0.576,), 10% (0.655), 5% (0.786), 2.5(0.9187)%, 1 (1.091)%  *Data is not normally distributed (reject H0) | Statistic: 245.83* > greater than the critical values:-  15% (0.576,), 10% (0.656), 5% (0.787), 2.5(0.787)%, 1 (1.092)%  *Data is not normally distributed (reject H0) |
| **Death** | Statistic: 60.48* > greater than the critical values:-  15% (0.576,), 10% (0.655), 5% (0.786), 2.5(0.9187)%, 1 (1.091)%  *Data is not normally distributed (reject H0) | Statistic: 222.75* > greater than the critical values:-  15% (0.576,), 10% (0.656), 5% (0.787), 2.5(0.787)%, 1 (1.092)%  *Data is not normally distributed (reject H0) |

### **S.4 difference between each NMPDU category and non-NMPDU (control group)**

#### **Emotional analysis**

Table 4 presents the median Mann–Whitney U test results and the effect size of the comparison between the users from the categories (i.e., opioids, benzodiazepines, stimulants, and polysubstance) of the NMPDU group and the users from the control group.

**Opioid NMPDU group versus control group:** As shown in Table 4, the users from the opioid NMPDU group significantly tend to use more words related to fear (p < 0.001, r = 0.25), anger (p < 0.001, r = 0.47), negative sentiment (p < 0.001, r = 0.37), sadness (p < 0.001, r = 0.27), and disgust (p < 0.001, r = 0.48) than the users from the control group. Opioid NMPDU users use significantly less words related to positive sentiment (p < 0.001, r = 0.34), joy (p < 0.001, r = 0.33), trust (p < 0.001, r = 0.24), anticipation (p < 0.001, r = 0.36), and surprise (p < 0.001, r = 0.33) than the users from the control group.

**Stimulant NMPDU group versus control group:** The users from the stimulant NMPDU group tend to use significantly more words related to fear (p < 0.001, r = 0.28), anger (p < 0.001, r = 0.52), negative sentiment (p < 0.001, r = 0.41), sadness (p < 0.001, r = 0.33), and disgust (p < 0.001, r = 0.56) than the users from the control group. Stimulant NMPDU users use significantly less words related to positive sentiment (p < 0.001, r = 0.38), joy (p < 0.001, r = 0.35), trust (p < 0.001, r = 0.25), anticipation (p < 0.001, r = 0.42), and surprise (p < 0.001, r = 0.36) than the users from the control group (Table 4).

**Benzodiazepine NMPDU group versus control group:** As presented in Table 4, the users from benzodiazepine NMPDU group significantly tend to use more words related to fear (p < 0.001, r = 0.34), anger (p < 0.001, r = 0.53), negative sentiment (p < 0.001, r = 0.44), sadness (p < 0.001, r = 0.39), and disgust (p < 0.001, r = 0.56) than the users from the control group. Benzodiazepine NMPDU users use significantly less words related to positive sentiment (p < 0.001, r = 0.29), joy (p < 0.001, r = 0.26), trust (p < 0.001, r = 0.190), anticipation (p < 0.001, r = 0.32), and surprise (p < 0.001, r = 0.29) than the users from the control group.

**Polysubstance NMPDU group versus control group:** The users from polysubstance NMPDU group significantly tends to use more words related to fear (p < 0.001, r = 0.33), anger (p < 0.001, r = 0.52), negative sentiment (p < 0.001, r = 0.43), sadness (p < 0.001, r = 0.38), and disgust (p < 0.001, r = 0.55) than the users from the control group. Polysubstance NMPDU users use significantly less words related to positive sentiment (p < 0.001, r = 0.32), joy (p < 0.001, r = 0.28), trust (p < 0.001, r = 0.21), anticipation (p < 0.001, r = 0.32), and surprise (p < 0.001, r = 0.29) than the users from the control group (Table 4)

Table 4 Comparison of emotional differences between users from the NMPDU and control groups

| **Drug category** | **Emotions** | **Median: NMPDU group** | **Median: Control group** | **Different (NMPDU versus Control)**  **Mann–Whitney U test results** | |
| --- | --- | --- | --- | --- | --- |
|  |  |  |  | **P-value (p)** | **Effect sizes (r)** |
| **Opioids** | **Positive** | **0.513** | **0.696** | **<0.001** | **0.34**** |
|  | **Trust** | **0.315** | **0.393** | **<0.001** | **0.24*** |
|  | **Anticipation** | **0.284** | **0.396** | **<0.001** | **0.36**** |
|  | **Fear** | **0.2192** | **0.176** | **<0.001** | **0.25*** |
|  | **Anger** | **0.250** | **0.148** | **<0.001** | **0.47**** |
|  | **Negative** | **0.468** | **0.337** | **<0.001** | **0.37**** |
|  | **Sadness** | **0.213** | **0.169** | **<0.001** | **0.27*** |
|  | **Joy** | **0.266** | **0.368** | **<0.001** | **0.33**** |
|  | **Surprise** | **0.138** | **0.190** | **<0.001** | **0.33**** |
|  | **Disgust** | **0.206** | **0.113** | **<0.001** | **0.48**** |
| **Stimulant** | **Positive** | **0.541** | **0.696** | **<0.001** | **0.38**** |
|  | **Trust** | **0.331** | **0.393** | **<0.001** | **0.25*** |
|  | **Anticipation** | **0.303** | **0.396** | **<0.001** | **0.42**** |
|  | **Fear** | **0.213** | **0.176** | **<0.001** | **0.28*** |
|  | **Anger** | **0.227** | **0.148** | **<0.001** | **0.52***** |
|  | **Negative** | **0.449** | **0.337** | **<0.001** | **0.41**** |
|  | **Sadness** | **0.212** | **0.169** | **<0.001** | **0.33*** |
|  | **Joy** | **0.287** | **0.368** | **<0.001** | **0.35**** |
|  | **Surprise** | **0.147** | **0.190** | **<0.001** | **0.36**** |
|  | **Disgust** | **0.191** | **0.113** | **<0.001** | **0.56***** |
| **Benzodiazepines** | **Positive** | **0.563** | **0.696** | **<0.001** | **0.29*** |
|  | **Trust** | **0.341** | **0.393** | **<0.001** | **0.19*** |
|  | **Anticipation** | **0.316** | **0.396** | **<0.001** | **0.32**** |
|  | **Fear** | **0.229** | **0.176** | **<0.001** | **0.34**** |
|  | **Anger** | **0.241** | **0.148** | **<0.001** | **0.53***** |
|  | **Negative** | **0.475** | **0.337** | **<0.001** | **0.44**** |
|  | **Sadness** | **0.227** | **0.169** | **<0.001** | **0.39**** |
|  | **Joy** | **0.303** | **0.368** | **<0.001** | **0.26*** |
|  | **Surprise** | **0.152** | **0.190** | **<0.001** | **0.29**** |
|  | **Disgust** | **0.202** | **0.113** | **<0.001** | **0.56***** |
| **Polysubstance** | **Positive** | **0.563** | **0.696** | **<0.001** | **0.29*** |
|  | **Trust** | **0.341** | **0.393** | **<0.001** | **0.190*** |
|  | **Anticipation** | **0.316** | **0.396** | **<0.001** | **0.32**** |
|  | **Fear** | **0.229** | **0.176** | **<0.001** | **0.34**** |
|  | **Anger** | **0.241** | **0.148** | **<0.001** | **0.53***** |
|  | **Negative** | **0.47** | **0.337** | **<0.001** | **0.44**** |
|  | **Sadness** | **0.227** | **0.169** | **<0.001** | **0.39**** |
|  | **Joy** | **0.303** | **0.368** | **<0.001** | **0.26*** |
|  | **Surprise** | **0.152** | **0.190** | **<0.001** | **0.29*** |
|  | **Disgust** | **0.202** | **0.113** | **<0.001** | **0.56***** |

*** Small difference (0.1 ≤ r < 0.3). **Medium difference (0.3 ≤ r < 0.5). ***Large difference (0.5 ≤ r < 0.7). ****Very large difference (r ≥ 0.7)**

#### **Personal and social concern analysis**

Table 5 presents the median Mann–Whitney U test results and the effect size of the comparison between the users from the categories (i.e., opioids, benzodiazepines, stimulants, and polysubstance) of the NMPDU group and the users from the control group.

**Opioid NMPDU group versus control group:** As shown in Table 3, the users from the opioid NMPDU group express significantly more social contents related to family (p < 0.001, r = 0.133) and less social contents related to friends (p < 0.001, r = 0.215) than the users from the control group. For personal concern contents, the users from the opioid NMPDU group express significantly less personal concern contents related to work (p < 0.001, r = 0.57), leisure (p < 0.001, r = 0.61), home (p < 0.001, r = 0.50), money (p < 0.001, r = 0.039), and religion (p < 0.001, r = 0.21) than the users from the control group, and no significant difference exists in the death variable (p > 0.001). Comparing both groups based on biological process contents demonstrates that the users from opioid NMPDU group intend to use less contents related to health (p < 0.001, r = 0.24) and use more contents related to the body (p < 0.001, r = 0.26) than the users from the control group. Comparing both groups based on time orientation content shows that the users from opioid NMPDU group intend to discuss significantly more contents related to the past (p < 0.001, r = 0.457) than the users from the control group. Finally, the users from the opioid NMPDU group express significantly less core drive contents related to achievement (p < 0.001, r = 0.56).

**Stimulant NMPDU group versus control group:** The users from the stimulant NMPDU group express significantly more social contents related to family (p < 0.001, r = 0.197) and no significant difference is observed in contents related to friends (p > 0.001) than the users from the control group (Table 6). The comparison between the two groups based on personal concern content demonstrates that the users from the stimulant NMPDU group express significantly less personal concern contents related to work (p < 0.001, r = 0.55), leisure (p < 0.001, r = 0.61, home (p < 0.001, r = 0.49), money (p < 0.001, r = 0.041), and religion (p < 0.001, r = 0.21) than the users from the control group. No significant difference is found in the death variable (p > 0.001). For biological process contents, the users from the stimulant NMPDU group intend to use less contents related to health (p < 0.001, r = 0.24) and use more contents related to the body (p < 0.001, r = 0.26) than the users from the control group. Comparing both groups based on time orientation contents shows that the users from the stimulant NMPDU group intend to discuss significantly more contents related to the past (p < 0.001, r = 0.457) than the users from the control group. Finally, the users from the stimulant NMPDU group express significantly less core drive contents related to achievement (p < 0.001, r = 0.62) than the users from the control group.

**Benzodiazepine NMPDU group versus control group:** The users from the benzodiazepine NMPDU group express significantly more social contents related to family (p < 0.001, r = 0.182) and less social contents related to friends (p < 0.001, r = 0.019) than the users from the control group. The users from the benzodiazepine NMPDU group express significantly less personal concern contents related to work (p < 0.001, r = 0.59), leisure (p < 0.001, r = 0.67, home (p < 0.001, r = 0.46), money (p < 0.001, r = 0.041), and religion (p < 0.001, r = 0.23) and express more contents related to death (p < 0.001, r = 0.097) than the users from the control group. For biological process contents, the users from the benzodiazepine NMPDU group intend to use less contents related to health (p < 0.001, r = 0.10) and use more contents related to the body (p < 0.001, r = 0.25) than the users from the control group. Comparing both groups based on time orientation contents shows that the users from benzodiazepine NMPDU group intend to discuss significantly more contents related to the past (p < 0.001, r = 0.55) than the users from the control group. Finally the users from benzodiazepine NMPDU group express significantly less core drive contents related to achievement (p < 0.001, r = 0.60) than the users from the control group (Table 3).

**Polysubstance NMPDU group versus control group:** As presented in Table 6, the users from the polysubstance NMPDU group express significantly more social contents related to family (p < 0.001, r = 0.197) and no significant difference is observed in contents related to friends (p > 0.001) than the users from the control group. For personal concern contents, the users from the polysubstance NMPDU group express significantly less contents related to work (p < 0.001, r = 0.51), leisure (p < 0.001, r = 0.58), home (p < 0.001, r = 0.38), money (p < 0.001, r = 0.030), and religion (p < 0.001, r = 0.144) and express significantly more contents related to death (p < 0.001, r = 0.18) than the users from the control group. Comparing both groups based on biological process contents demonstrates that the users from the polysubstance NMPDU group have no significant difference in using health-related contents (p > 0.001) but use more contents related to the body (p < 0.001, r = 0.35) than the users from the control group. Time orientation related contents show that the users from the polysubstance NMPDU group intend to discuss significantly more contents related to the past (p < 0.001, r = 0.457) than the users from the control group. Finally, users from the polysubstance NMPDU group express significantly less core drive contents related to achievement (p < 0.001, r = 0.54) than the users from the control group.

Table 5 Comparing personal and social differences between the users from the NMPDU and control groups

| **Drug category** | **Content** | **Variables** | **Median: NMPDU group** | **Median: Control group** | **Different (NMPDU versus control)**  **Mann–Whitney U test results** | |
| --- | --- | --- | --- | --- | --- | --- |
|  |  |  |  |  | **P-value (p)** | **Effect sizes (r)** |
| **Opioids** | **Social**  **content** | **Family** | **0.0319** | **0.025** | **<0.001** | **0.133*** |
|  |  | **Friend** | **0.013** | **0.020** | **<0.001** | **0.215*** |
|  | **Time orientation**  **content** | **Past focus** | **0.231** | **0.140** | **<0.001** | **0.457**** |
|  | **Core drive content** | **Achieve** | **0.092** | **0.204** | **<0.001** | **0.56***** |
|  | **Biological process**  **content** | **Health** | **0.041** | **0.056** | **<0.001** | **0.24*** |
|  |  | **Body** | **0.117** | **0.086** | **<0.001** | **0.26*** |
|  | **Personal concern**  **content** | **Work** | **0.075** | **0.215** | **<0.001** | **0.57***** |
|  |  | **Leisure** | **0.103** | **0.272** | **<0.001** | **0.61***** |
|  |  | **Home** | **0.025** | **0.058** | **<0.001** | **0.50***** |
|  |  | **Money** | **0.051** | **0.086** | **<0.001** | **0.39**** |
|  |  | **Religion** | **0.032** | **0.041** | **>0.001** | **0.21*** |
|  |  | **Death** | **0.016** | **0.016** | **>0.001** | **-** |
| **Stimulant** | **Social**  **content** | **Family** | **0.033** | **0.025** | **<0.001** | **0.197*** |
|  |  | **Friend** | **0.019** | **0.020** | **>0.001** | **-** |
|  | **Time orientation content** | **Past focus** | **0.240** | **0.140** | **<0.001** | **0.60***** |
|  | **Core drive content** | **Achieve** | **0.111** | **0.204** | **<0.001** | **0.62***** |
|  | **Biological process content** | **Health** | **0.046** | **0.056** | **<0.001** | **-** |
|  |  | **Body** | **0.105** | **0.086** | **<0.001** | **0.22*** |
|  | **Personal concern content** | **Work** | **0.111** | **0.215** | **<0.001** | **0.55***** |
|  |  | **Leisure** | **0.123** | **0.272** | **<0.001** | **0.61***** |
|  |  | **Home** | **0.032** | **0.058** | **<0.001** | **0.49***** |
|  |  | **Money** | **0.055** | **0.086** | **<0.001** | **0.41**** |
|  |  | **Religion** | **0.032** | **0.041** | **>0.001** | **0.21*** |
|  |  | **Death** | **0.017** | **0.016** | **>0.001** | **-** |
| **Benzodiazepines** | **Social**  **content** | **Family** | **0.033** | **0.025** | **<0.001** | **0.182*** |
|  |  | **Friend** | **0.019** | **0.020** | **<0.001** | **0.019*** |
|  | **Time orientation content** | **Past focus** | **0.246** | **0.140** | **<0.001** | **0.55**** |
|  | **Core drive content** | **Achieve** | **0.110** | **0.204** | **<0.001** | **0.60***** |
|  | **Biological process content** | **Health** | **0.050** | **0.056** | **<0.001** | **0.10*** |
|  |  | **Body** | **0.112** | **0.086** | **<0.001** | **0.25*** |
|  | **Personal concern content** | **Work** | **0.097** | **0.215** | **<0.001** | **0.59***** |
|  |  | **Leisure** | **0.123** | **0.272** | **<0.001** | **0.67**** |
|  |  | **Home** | **0.032** | **0.058** | **<0.001** | **0.46**** |
|  |  | **Money** | **0.055** | **0.086** | **<0.001** | **0.41**** |
|  |  | **Religion** | **0.032** | **0.041** | **<0.001** | **0.23*** |
|  |  | **Death** | **0.018** | **0.016** | **<0.001** | **0.097** |
| **Polysubstance** | **Social**  **content** | **Family** | **0.035** | **0.025** | **<0.001** | **0.197*** |
|  |  | **Friend** | **0.019** | **0.020** | **>0.001** | **-** |
|  | **Time orientation content** | **Past focus** | **0.254** | **0.140** | **<0.001** | **0.53**** |
|  | **Core drive content** | **Achieve** | **0.107** | **0.204** | **<0.001** | **0.54***** |
|  | **Biological process content** | **Health** | **0.056** | **0.056** | **>0.001** | **-** |
|  |  | **Body** | **0.128** | **0.086** | **<0.001** | **0.35**** |
|  | **Personal concern content** | **Work** | **0.096** | **0.215** | **<0.001** | **0.51***** |
|  |  | **Leisure** | **0.125** | **0.272** | **<0.001** | **0.58**** |
|  |  | **Home** | **0.033** | **0.058** | **<0.001** | **0.38**** |
|  |  | **Money** | **0.059** | **0.086** | **<0.001** | **0.30**** |
|  |  | **Religion** | **0.035** | **0.041** | **<0.001** | **0.144*** |
|  |  | **Death** | **0.020** | **0.016** | **<0.001** | **0.18*** |

*** Small difference (0.1 ≤ r < 0.3). **Medium difference (0.3 ≤ r < 0.5). ***Large difference (0.5 ≤ r < 0.7). ****Very large difference (r ≥ 0.7)**

### **S.5 Topic modeling**

We then inspected the word clusters in each set of sub-topics that can give the most useful topics. Twenty of the frequently used words for the top 10 sub-topics related to each category are presented in the table below.

***Supplementary Table 6 Opioid (Top 20 words across each detected sub-topic-related topic by LDA)***

| ***Top 20 words across each 10 detected sub-topic related topic by LDA*** | ***Manual topic classification*** |
| --- | --- |
| ***Topic: 0***  shit, high, popped, took, weed, left, addict, buzz, caught, away, Hooked and addiction , back, wish, take, stay, really, days, help, couple, send, popping, teeth, brother, relax, work, cool, ibuprofen, brought, smoke, know  ***Topic: 1***  high, like, feel, popped, take, hell, taking, batman, moon, addicted, wanna, bout, trip, extra, drugs, takin, weed, pack, body, money, dope, took, nauseous, make, original, quarter, pass, coke, shit, list  ***Topic: 2***  another, wine, sleep, smoking, molly, took, high, like, remember, mixing, time, taking, even, lean, going, coffee, wake, dirty, drink, shot, gonna, good, bottle, take, might, ibuprofen, years, know, drank, drinking  ***Topic: 3***  high, drunk, getting, eating, dose, strippers, stress, ease, good, time, like, selling, best, took, years, really, weed, life, today, surgery, gotta, liquor, demons, sure, whole, recreational, picked, sister, lean, first  ***Topic: 4***  need, high, popped, enough, someone, please, bought, think, feelings, make, help, still, getting, pain, tired, thank, took, addiction, house, come, friends, blunt, much, give, gettin, crush, life, seen, bring, weed  ***Topic: 5***  like, night, pill, last, going, high, lean, right, took, bottle, snort, done, shit, half, things, take, gave, tough, wants, pain, back, knock, feeling, pretty, good, popping, hours, trade, sold, turn  ***Topic: 6***  high, joint, love, work, baby, keep, drop, lmao, think, night, took, time, shit, came, hours, tryna, found, know, alcohol, damn, sell, sleep, drugs, swear, pour, overdose, still, floor, last, hard  ***Topic: 7***  like, time, dude, geeked, smart, whiskey, plug, straight, much, never, touch, days, bull, beer, high, felt, paris, today, gonna, glass, smoking, sniffing, rules, call, hour, floating, hydro, full, heroin, drinking  ***Topic: 8***  want, pills, pain, find, anyone, might, night, much, popping, cause, know, kill, took, vodka, take, weed, stop, back, found, right, make, could, sell, poppin, couple, mixed, even, cant, tell, shit  ***Topic: 9***  give, still, friend, gave, high, like, took, could, take, homie, wanted, asking, know, looking, anyone, asked, today, back, year, bring, shit, come, bottle, traded, problems, really, wanna, cheap, itchin, itching | - To relieve physical pain (e.g. pain, hospital, nauseous, surgery) - To help with sleep (e.g. sleep) - To get high or use it with illicit drug (e.g. high, dope , heroin) - To help with emotions(e.g. stress) - Hooked and addiction (e.g. hook , addiction) - To relax (e.g. relax, cool, recreational, good) - To use it with smoking e.g. (smoking ,blunt, weed) - To use with alcohol (e.g. drunk, liquor, wine, whiskey, vodka) |

***Supplementary Table 7 Stimulants (Top 20 words across each detected sub-topic related topic by LDA)***

| ***Top 20 words across each 10 detected sub-topic related topic by LDA*** | ***Manual topic classification*** |
| --- | --- |
| Topic: 0  much, snort, weed, high, plug, snorted, alcohol, need, right, today, sniff, line, could, make, lines, another, really, take, like, happens, wish, morning, know, crush, many, school, smoking, crushed, rail, little  Topic: 1  paper, took, take, write, tomorrow, finish, final, homework, essay, night, might, class, wanna, popped, page, done, semester, college, work, study, like, trying, hours, hour, writing, exam, around, help, thank, coffee  Topic: 2  took, like, time, today, study, feel, want, accidentally, first, instead, enough, much, know, morning, asleep, addicted, said, good, test, shit, take, takes, tried, never, went, still, could, finals, college, think  Topic: 3  coffee, friend, asking, today, breakfast, espresso, like, iced, gave, extra, shots, wait, shot, stop, energy, selling, drinks, told, going, idea, snorts, hydro, best, large, caffeine, good, monster, morning, doctor, feeling  Topic: 4  coffee, like, bull, take, drink, feel, cold, caffeine, cups, gotta, today, brew, shit, heart, tired, drinking, drank, need, night, redbull, days, morning, nothing, four, crack, going, taking, love, either, body  Topic: 5  snorting, need, gonna, work, take, someone, like, shit, clean, done, help, focus, give, room, know, really, homework, think, addiction, today, friends, start, tonight, house, study, school, even, bring, make, going  Topic: 6  time, anyone, glass, whiskey, jeans, diesel, shower, someone, please, sell, know, anybody, smile, somebody, plug, wants, needs, wine, hook, find, send, sells, give, knows, wanna, girl, tryna, connect, could, want  Topic: 7  hours, sleep, took, night, last, days, like, time, work, studying, take, year, study, still, straight, three, taking, finals, haven, hour, much, awake, week, half, coffee, stay, first, today, exam, slept  Topic: 8  week, finals, taking, diet, lean, mixing, back, wake, coke, coffee, sleep, need, drinking, time, college, going, school, coming, shit, like, next, stay, think, really, want, semester, well, miss, caffeine, high  Topic: 9  crushed, sure, good, sniffing, pretty, morning, dose, found, double, high, ready, someone, left, coke, time, hitting, popping, mixed, full, yeah, ever, college, snorted, remember, work, never, diet, addiction, supply, mine | - To help study(e.g. finals, college, semester, study, school, exam, writing, help , test, homework, essay) - To stay awake (e.g , awake, espresso redbull, caffeine) - To use with alcohol (e.g. drank, wine, whiskey) - Hooked and addiction (e.g. hook , addiction) - To use it with smoking e.g. smoking , weed) |

***Supplementary Table 8 Benzodiazepines (Top 20 words across each detected sub-topic related topic by LDA)***

| ***Top 20 words across each 10 detected sub-topic related topic by LDA*** | ***Manual topic classification*** |
| --- | --- |
| Topic: 0  like, feel, want, addiction, know, good, alcohol, take, sell, high, weed, today, really, still, life, going, pills, months, better, took, make, need, feeling, days, anxiety, tryna, right, gonna, well, happy  Topic: 1  take, popped, another, love, high, wanna, time, like, going, fake, gave, coke, half, know, whiskey, happens, shit, damaged, bars, molly, night, tequila, glass, life, much, year, blow, four, handful, right  Topic: 2  weed, work, take, smoke, tonight, need, gonna, enough, plug, high, smoking, lean, really, blunt, make, wait, today, trying, coffee, help, alcohol, drinking, diet, going, right, days, back, school, drink, coke  Topic: 3  time, like, last, night, think, take, bottle, wine, first, high, drugs, year, weed, remember, much, used, taking, alcohol, lots, week, self, life, still, took, really, snorting, cause, drink, lmao, even  Topic: 4  anyone, done, addicted, time, asleep, shit, year, anybody, even, half, took, today, still, life, around, like, know, last, ever, never, going, wanna, yeah, give, face, chill, haven, hours, taken, hour  Topic: 5  took, sleep, found, back, shit, went, today, time, first, party, rest, dead, going, might, cold, peace, could, miss, good, many, head, smile, accidentally, bunch, hand, wish, little, girl, brew, extra  Topic: 6  much, bars, could, never, sorry, cocaine, years, beer, still, know, remember, need, bout, thought, clean, share, home, liquor, tell, poppin, taking, somebody, getting, tonight, alcohol, slip, talking, asked, like, give  Topic: 7  someone, please, bring, send, give, anyone, vodka, head, marijuana, call, shit, strongest, shots, mixed, find, hear, know, friends, cool, dude, today, miss, wants, come, gonna, need, snort, pass, want, voices  Topic: 8  wine, night, took, last, morning, hours, tomorrow, work, coffee, sleep, woke, bottle, glass, wake, drank, today, like, much, later, taking, good, slept, vodka, breakfast, still, going, next, beer, gave, whole  Topic: 9  need, friend, coffee, take, asking, time, gonna, popping, sleep, like, much, flight, caffeine, high, couple, right, good, three, everything, best, ever, really, taking, today, attack, going, know, panic, days, something | - To use with alcohol (e.g. drank, vodka, tequila, liquor, wine, whiskey) - To get high or use it with illicit drug (e.g. high, cocaine) - To help with sleep (e.g. sleep, asleep, slept) - To help with emotions(e.g. anxiety ) - Hooked and addiction (e.g. addiction) - To relax (e.g. relax, cool, recreational, happy) - To use it with smoking e.g. (smoking , marijuana ,blunt, weed) |

***Supplementary Table 9 Polysubstance (Top 20 words across each detected sub-topic related topic by LDA)***

| ***Polysubstance (Top 20 words across each detected Stimulants related topic by LDA)*** | ***Manual topic classification*** |
| --- | --- |
| Topic: 0  took, like, high, night, sleep, time, hours, take, shit, last, taking, good, back, work, feel, still, went, today, first, make, gonna, going, bunch, hour, life, days, drinking, found, much, hell  Topic: 1  want, lean, taking, wake, week, mixing, finals, friend, know, asking, wine, eating, going, sleep, take, study, hydro, like, forget, give, think, anyone, someone, next, feeling, hope, days, dinner, girl, birthday  Topic: 2  wanna, take, many, like, drunk, took, please, plug, someone, night, bars, really, friends, good, send, kinda, getting, gonna, chill, happens, right, weed, kill, much, snorting, molly, think, found, pharmacy, life  Topic: 3  weed, money, tonight, nothing, home, looking, wish, paper, love, anybody, like, gonna, took, know, sleep, gave, good, week, high, made, years, ecstasy, wanted, wants, pussy, world, faded, could, combo, popped  Topic: 4  much, took, feel, found, like, snort, today, dose, time, glass, shit, whiskey, take, head, half, heart, popped, lmao, back, stay, could, shower, joint, strippers, cold, jeans, drank, away, think, smoke  Topic: 5  need, like, year, sell, someone, high, popping, drugs, used, gave, really, back, weed, years, addicted, know, shit, time, alcohol, take, sober, last, months, coke, still, since, addiction, times, bring, cocaine  Topic: 6  take, love, popped, need, still, tomorrow, another, gonna, weed, even, coke, drink, getting, morning, coffee, addicted, done, like, shots, going, today, never, smoke, work, gotta, ever, shit, vodka, sleep, ready  Topic: 7  coffee, time, shit, today, never, remember, took, days, addiction, hours, blunt, molly, addict, breakfast, iced, work, night, drinking, back, anything, even, good, last, take, literally, self, felt, mixed, cups, friend  Topic: 8  prescription, online, order, snorting, cheap, might, without, sale, high, addiction, concerta, poppin, call, quality, start, discount, body, really, required, cigarettes, heroin, perc, drug, trying, around, wattpad, friend, half, couple, friends  Topic: 9  find, best, anyone, pills, free, need, right, make, online, generic, cheap, shipping, satisfaction, could, extra, work, house, night, cleaning, room, bought, come, yeah, feelings, help, know, tonight, friend, clean, friends | - To help study(e.g. finals, study) - To get high or use it with illicit drug (e.g. high, heroin, cocaine) - To help with emotions(e.g. ecstasy, satisfaction) - Hooked and addiction ( addiction , addict) - To socialize (birthday ,couple, friends) - To use it with smoking e.g. (cigarettes, blunt, weed) - To use with alcohol (e.g. drunk, wine, whiskey, vodka) - To help with sleep (e.g. sleep) |

### **S.6 References**

[6]	R. N. Lipari, M. Williams, and S. L. Van Horn, "Why do adults misuse prescription drugs?," in *The CBHSQ Report*: Substance Abuse and Mental Health Services Administration (US), 2017.

[7]	T. W. Anderson and D. A. Darling, "A test of goodness of fit," *Journal of the American statistical association,* vol. 49, no. 268, pp. 765-769, 1954.

[8]	S. Engmann and D. Cousineau, "Comparing distributions: the two-sample Anderson-Darling test as an alternative to the Kolmogorov-Smirnoff test," *Journal of applied quantitative methods,* vol. 6, no. 3, pp. 1-17, 2011.

[9]	R. B. D'Agostino, *Goodness-of-fit-techniques*. CRC press, 1986.
